# Protection afforded by prior infection against SARS-CoV-2 reinfection with the Omicron variant

**DOI:** 10.1101/2022.01.05.22268782

**Authors:** Heba Altarawneh, Hiam Chemaitelly, Patrick Tang, Mohammad R. Hasan, Suelen Qassim, Houssein H. Ayoub, Sawsan AlMukdad, Hadi M. Yassine, Fatiha M. Benslimane, Hebah A. Al Khatib, Peter Coyle, Zaina Al Kanaani, Einas Al Kuwari, Andrew Jeremijenko, Anvar Hassan Kaleeckal, Ali Nizar Latif, Riyazuddin Mohammad Shaik, Hanan F. Abdul Rahim, Gheyath K. Nasrallah, Mohamed Ghaith Al Kuwari, Adeel A. Butt, Hamad Eid Al Romaihi, Mohamed H. Al-Thani, Abdullatif Al Khal, Roberto Bertollini, Laith J. Abu-Raddad

**Affiliations:** Infectious Disease Epidemiology Group, Weill Cornell Medicine-Qatar, Cornell University, Doha, Qatar; World Health Organization Collaborating Centre for Disease Epidemiology Analytics on HIV/AIDS, Sexually Transmitted Infections, and Viral Hepatitis, Weill Cornell Medicine–Qatar, Cornell University, Qatar Foundation – Education City, Doha, Qatar; Department of Pathology, Sidra Medicine, Doha, Qatar; Mathematics Program, Department of Mathematics, Statistics, and Physics, College of Arts and Sciences, Qatar University, Doha, Qatar; Biomedical Research Center, Member of QU Health, Qatar University, Doha, Qatar; Department of Biomedical Science, College of Health Sciences, Member of QU Health, Qatar University, Doha, Qatar; Hamad Medical Corporation, Doha, Qatar; Wellcome-Wolfson Institute for Experimental Medicine, Queens University, Belfast, United Kingdom; College of Health Sciences, QU Health, Qatar University, Doha, Qatar; Primary Health Care Corporation, Doha, Qatar; Department of Population Health Sciences, Weill Cornell Medicine, Cornell University, New York, New York, USA; Ministry of Public Health, Doha, Qatar; Department of Public Health, College of Health Sciences, Member of QU Health, Qatar University, Doha, Qatar

## Abstract

**BACKGROUND:** Natural SARS-CoV-2 infection elicits strong protection against reinfection with the Alpha (B.1.1.7), Beta (B.1.351), and Delta (B.1.617.2) variants. However, the Omicron (B.1.1.529) variant harbors multiple mutations that can mediate immune evasion. We estimated effectiveness of prior infection in preventing reinfection (*PES*) with Omicron and other SARS-CoV-2 variants in Qatar.

**METHODS:** *PES* was estimated using the test-negative, case-control study design, employing a methodology that was recently investigated and validated for derivation of robust estimates for *PES*. Cases (PCR-positive persons with a variant infection) and controls (PCR-negative persons) were exact-matched by sex, 10-year age group, nationality, and calendar time of PCR test, to control for known differences in the risk of exposure to SARS-CoV-2 infection in Qatar.

**RESULTS:** *PES* against symptomatic reinfection was estimated at 90.2% (95% CI: 60.2-97.6) for Alpha, 84.8% (95% CI: 74.5-91.0) for Beta, 92.0% (95% CI: 87.9-94.7) for Delta, and 56.0% (95% CI: 50.6-60.9) for Omicron. Only 1 Alpha, 2 Beta, 0 Delta, and 2 Omicron reinfections progressed to severe COVID-19. None progressed to critical or fatal COVID-19. *PES* against hospitalization or death due to reinfection was estimated at 69.4% (95% CI: −143.6-96.2) for Alpha, 88.0% (95% CI: 50.7-97.1) for Beta, 100% (95% CI: 43.3-99.8) for Delta, and 87.8% (95% CI: 47.5-97.1) for Omicron.

**CONCLUSIONS:** Protection afforded by prior infection in preventing symptomatic reinfection with Alpha, Beta, or Delta is robust, at about 90%. While such protection against reinfection with Omicron is lower, it is still considerable at nearly 60%. Prior-infection protection against hospitalization or death at reinfection appears robust, regardless of variant.

## Introduction

Natural severe acute respiratory syndrome coronavirus 2 (SARS-CoV-2) infection elicits strong protection against reinfection with the Alpha (B.1.1.7),^1,2^ Beta (B.1.351),^1^ and Delta (B.1.617.2)^3^ variants. However, the Omicron (B.1.1.529) variant harbors multiple mutations that can mediate immune evasion. We estimated effectiveness of prior infection in preventing reinfection (*PES*) with Omicron and other SARS-CoV-2 variants in Qatar.

## Methods

### Study population, data sources, and study design

This study was conducted in the resident population of Qatar, applying the test-negative, case-control study design^4–6^ to investigate the protection afforded by prior SARS-CoV-2 infection in preventing reinfection with SARS-CoV-2 variants. Effectiveness of prior infection in preventing reinfection (*PES*) was defined as the proportional reduction in susceptibility to infection among those with prior infection versus those without.^6,7^ The test-negative methodology was recently investigated and validated for the specific derivation of rigorous and robust estimates for SARS-CoV-2 *PES*.^6^

Coronavirus Disease 2019 (COVID-19) laboratory testing, vaccination, clinical infection data, and related demographic details were extracted from the national, federated SARS-CoV-2 databases that include all polymerase chain reaction (PCR) testing, COVID-19 vaccinations, and COVID-19 hospitalizations and deaths in Qatar since the start of the pandemic, with no missing information on variables included in this study.

Every PCR test conducted in Qatar is classified based on the reason for testing (clinical symptoms, contact tracing, surveys or random testing campaigns, individual requests, routine healthcare testing, pre-travel, at port of entry, or other). Qatar has unusually young, diverse demographics, in that only 9% of its residents are ≥50 years of age, and 89% are expatriates from over 150 countries.^8,9^ Nearly all individuals were vaccinated in Qatar, however, vaccinations performed elsewhere were still recorded in the health system at the port of entry upon arrival to Qatar per country requirements.

For estimation of *PES* against the Alpha^10^ (B.1.1.7), Beta^10^ (B.1.351), and Delta^10^ (B.1.617.2) variants, cases (PCR-positive persons with genotyped variant infection) and controls (PCR-negative persons) identified between March 23, 2021 (start of positive samples’ genotyping in Qatar) and November 18, 2021 (prior to suspected introduction of the Omicron variant), were exact matched in a ratio of one-to-five by sex, 10-year age group, nationality, and calendar week of the PCR test (Figure 1 and Table 1). Infection with Alpha, Beta, or Delta variants was ascertained using real-time reverse-transcription PCR (RT-qPCR) genotyping of the positive clinical samples (Section S1).^11,12^

**Table 1.** Characteristics of matched cases (PCR-positive persons with Alpha, Beta, or Delta infections, respectively) and controls (PCR-negative persons).

| Characteristics | Cases*<br>(PCR-confirmed<br>infection with Alpha) | Controls*<br>(PCR-negative) | SMD<sup>§</sup> | Cases*<br>(PCR-confirmed<br>infection with Beta) | Controls*<br>(PCR-negative) | SMD<sup>§</sup> | Cases*<br>(PCR-confirmed<br>infection with Delta) | Controls*<br>(PCR-negative) | SMD<sup>§</sup> |
| --- | --- | --- | --- | --- | --- | --- | --- | --- | --- |
|  | N=336 | N=1,642 |  | N=1,336 | N=6,534 |  | N=2,176 | N=9,936 |  |
| **Median age (IQR) — years** | 31 (23-39) | 31 (23-39) | 0.00<sup>‡</sup> | 35 (27-42) | 34 (27-42) | 0.01<sup>‡</sup> | 31 (20-40) | 31 (19-40) | 0.04<sup>‡</sup> |
| **Age group — no. (%)** |  |  |  |  |  |  |  |  |  |
| <20 years | 70 (20.8) | 333 (20.3) |  | 157 (11.8) | 745 (11.4) |  | 538 (24.7) | 2,484 (25.0) |  |
| 20-29 years | 75 (22.3) | 372 (22.7) |  | 272 (20.4) | 1,346 (20.6) |  | 436 (20.0) | 2,056 (20.7) |  |
| 30-39 years | 113 (33.6) | 560 (34.1) |  | 471 (35.3) | 2,323 (35.6) |  | 615 (28.3) | 2,904 (29.2) |  |
| 40-49 years | 56 (16.7) | 267 (16.3) | 0.02 | 311 (23.3) | 1,522 (23.3) | 0.04 | 373 (17.1) | 1,626 (16.4) | 0.05 |
| 50-59 years | 18 (5.4) | 90 (5.5) |  | 97 (7.3) | 477 (7.3) |  | 153 (7.0) | 635 (6.4) |  |
| 60-69 years | 3 (0.9) | 15 (0.9) |  | 24 (1.8) | 112 (1.7) |  | 47 (2.2) | 174 (1.8) |  |
| 70+ years | 1 (0.3) | 5 (0.3) |  | 4 (0.3) | 9 (0.1) |  | 14 (0.6) | 53 (0.5) |  |
| **Sex** |  |  |  |  |  |  |  |  |  |
| Male | 178 (53.0) | 861 (52.4) | 0.01 | 937 (70.1) | 4,626 (70.8) | 0.01 | 1,108 (50.9) | 5,077 (51.1) | 0.00 |
| Female | 158 (47.0) | 781 (47.6) |  | 399 (29.9) | 1,908 (29.2) |  | 1,068 (49.1) | 4,859 (48.9) |  |
| **Nationality<sup>†</sup>** |  |  |  |  |  |  |  |  |  |
| Bangladeshi | 21 (6.3) | 105 (6.4) |  | 117 (8.8) | 581 (8.9) |  | 129 (5.9) | 621 (6.3) |  |
| Egyptian | 19 (5.7) | 95 (5.8) |  | 71 (5.3) | 351 (5.4) |  | 160 (7.4) | 718 (7.2) |  |
| Filipino | 43 (12.8) | 215 (13.1) |  | 132 (9.9) | 655 (10.0) |  | 175 (8.0) | 866 (8.7) |  |
| Indian | 55 (16.4) | 275 (16.7) |  | 309 (23.1) | 1,545 (23.6) |  | 202 (9.3) | 1,004 (10.1) |  |
| Nepalese | 17 (5.1) | 85 (5.2) | 0.04 | 163 (12.2) | 806 (12.3) | 0.04 | 38 (1.7) | 184 (1.9) | 0.12 |
| Pakistani | 13 (3.9) | 63 (3.8) |  | 55 (4.1) | 269 (4.1) |  | 67 (3.1) | 324 (3.3) |  |
| Qatari | 89 (26.5) | 445 (27.1) |  | 186 (13.9) | 930 (14.2) |  | 846 (38.9) | 4,127 (41.5) |  |
| Sri Lankan | 16 (4.8) | 78 (4.8) |  | 62 (4.6) | 301 (4.6) |  | 28 (1.3) | 130 (1.3) |  |
| Sudanese | 8 (2.4) | 36 (2.2) |  | 40 (3.0) | 197 (3.0) |  | 68 (3.1) | 280 (2.8) |  |
| Other nationalities<sup>‡</sup> | 55 (16.4) | 245 (14.9) |  | 201 (15.0) | 899 (13.8) |  | 463 (21.3) | 1,682 (16.9) |  |
| **PCR test calendar month\*\*** |  |  |  |  |  |  |  |  |  |
| March | 51 (15.2) | 349 (21.3) |  | 213 (15.9) | 1,375 (21.0) |  | 2 (0.1) | 10 (0.1) |  |
| April | 227 (67.6) | 1,018 (62.0) |  | 970 (72.6) | 4,417 (67.6) |  | 14 (0.6) | 67 (0.7) |  |
| May | 29 (8.6) | 130 (7.9) |  | 102 (7.6) | 497 (7.6) |  | 104 (4.8) | 495 (5.0) |  |
| June | 11 (3.3) | 50 (3.0) |  | 3 (0.2) | 23 (0.4) |  | 82 (3.8) | 409 (4.1) |  |
| July | 12 (3.6) | 78 (4.8) | 0.18 | 24 (1.8) | 106 (1.6) | 0.14 | 538 (24.7) | 2,384 (24.0) | 0.03 |
| August | 6 (1.8) | 17 (1.0) |  | 24 (1.8) | 116 (1.8) |  | 800 (36.8) | 3,704 (37.3) |  |
| September | 0 (0.0) | 0 (0.0) |  | 0 (0.0) | 0 (0.0) |  | 370 (17.0) | 1,678 (16.9) |  |
| October | 0 (0.0) | 0 (0.0) |  | 0 (0.0) | 0 (0.0) |  | 217 (10.0) | 977 (9.8) |  |
| November | 0 (0.0) | 0 (0.0) |  | 0 (0.0) | 0 (0.0) |  | 49 (2.3) | 212 (2.1) |  |

Abbreviations: IQR, interquartile range; PCR, polymerase chain reaction; SMD, standardized mean difference.
<sup>†</sup>Cases and controls were exact matched one-to-five by sex, 10-year age group, nationality, and calendar week of PCR test.
<sup>‡</sup>Nationalities were chosen to represent the most populous groups in Qatar.
<sup>§</sup>These comprise 20 other nationalities in Qatar in the Alpha variant analysis, 29 other nationalities in the Beta variant analysis, and 30 other nationalities in the Delta variant analysis.
<sup>§</sup>SMD is the difference in the mean of a covariate between groups divided by the pooled standard deviation. An SMD<0.1 indicates optimal balance in matching.
<sup>§</sup>SMD is for the mean difference between groups divided by the pooled standard deviation.
\*\*Cases and controls were exact matched using calendar week of PCR test, but we opted to report the distribution by calendar month for brevity. Accordingly, some cases and controls who were tested in the same week may appear in different calendar months.

**Figure 1.**
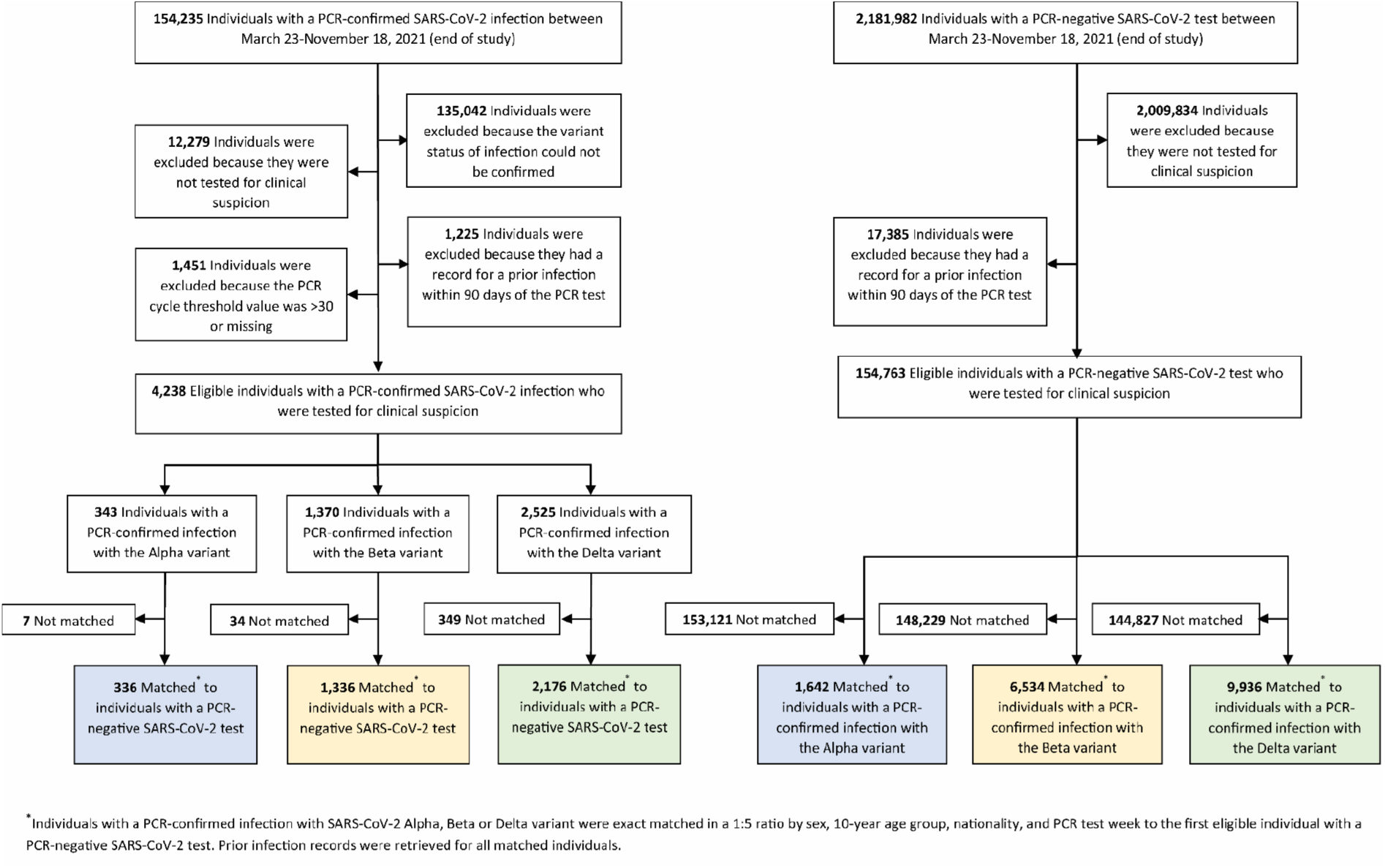
Flowchart describing the population selection process for investigating the effectiveness of prior infection in preventing reinfection with the Alpha, Beta, and Delta variants.

A similar methodology was applied to estimate *PES* against the Omicron^10^ (B.1.1.529) variant. However, cases (PCR-positive persons with Omicron infection) and controls (PCR-negative persons) identified between December 23 and January 2, 2022, the time during which the Omicron epidemic wave was exponentially growing in Qatar, were exact matched in a ratio of one-to-three by sex, 10-year age group, nationality, and calendar day of the PCR test (rather than calendar week of the PCR test; Figure 2 and Table 2). A SARS-CoV-2 infection with the Omicron variant was proxied as an S-gene “target failure” case using the TaqPath COVID-19 Combo Kit platform (Thermo Fisher Scientific, USA^13^) applying the criterion of an RT-qPCR Ct value ≤30 for both the N and ORF1ab genes, but a negative outcome for the S gene.

**Table 2.** Characteristics of matched cases (PCR-positive persons with Omicron infection) and controls (PCR-negative persons).

| Characteristics | Cases*<br>(PCR-confirmed infection with<br>Omicron) | Controls*<br>(PCR-negative) | SMD<sup>§</sup> |
| --- | --- | --- | --- |
|  | N=5,696 | N=10,673 |  |
| **Median age (IQR) — years** | 33 (25-40) | 32 (24-40) | 0.04<sup>†</sup> |
| **Age group — no. (%)** |  |  |  |
| <20 years | 923 (16.2) | 1,876 (17.6) | 0.06 |
| 20-29 years | 1,187 (20.8) | 2,339 (21.9) |  |
| 30-39 years | 2,078 (36.5) | 3,721 (34.9) |  |
| 40-49 years | 975 (17.1) | 1,718 (16.1) |  |
| 50-59 years | 369 (6.5) | 707 (6.6) |  |
| 60-69 years | 118 (2.1) | 232 (2.2) |  |
| 70+ years | 46 (0.8) | 80 (0.8) |  |
| **Sex** |  |  | 0.00 |
| Male | 3,148 (55.3) | 5,877 (55.1) |  |
| Female | 2,548 (44.7) | 4,796 (44.9) |  |
| **Nationality<sup>†</sup>** |  |  | 0.18 |
| Bangladeshi | 157 (2.8) | 323 (3.0) |  |
| Egyptian | 476 (8.4) | 746 (7.0) |  |
| Filipino | 1,003 (17.6) | 1,569 (14.7) |  |
| Indian | 1,027 (18.0) | 1,880 (17.6) |  |
| Nepalese | 170 (3.0) | 219 (2.1) |  |
| Pakistani | 160 (2.8) | 307 (2.9) |  |
| Qatari | 1,276 (22.4) | 3,126 (29.3) |  |
| Sri Lankan | 86 (1.5) | 122 (1.1) |  |
| Sudanese | 274 (4.8) | 550 (5.2) |  |
| Other nationalities<sup>‡</sup> | 1,067 (18.7) | 1,831 (17.2) |  |
| **PCR test date** |  |  | 0.13 |
| 23 December, 2021 | 244 (4.3) | 575 (5.4) |  |
| 24 December, 2021 | 127 (2.2) | 309 (2.9) |  |
| 25 December, 2021 | 287 (5.0) | 605 (5.7) |  |
| 26 December, 2021 | 540 (9.5) | 1,123 (10.5) |  |
| 27 December, 2021 | 657 (11.5) | 1,377 (12.9) |  |
| 28 December, 2021 | 887 (15.6) | 1,585 (14.9) |  |
| 29 December, 2021 | 1,043 (18.3) | 1,807 (16.9) |  |
| 30 December, 2021 | 1,199 (21.1) | 1,879 (17.6) |  |
| 31 December, 2021 | 670 (11.8) | 1,292 (12.1) |  |
| 01 January, 2022 | 42 (0.7) | 121 (1.1) |  |

Abbreviations: IQR, interquartile range; PCR, polymerase chain reaction; SMD, standardized mean difference.
\*Cases and controls were matched one-to-three by sex, 10-year age group, nationality, and PCR test date.
<sup>†</sup>Nationalities were chosen to represent the most populous groups in Qatar.
<sup>‡</sup>These comprise 44 other nationalities in Qatar.
<sup>§</sup>SMD is the difference in the mean of a covariate between groups divided by the pooled standard deviation. An SMD<0.1 indicates adequate matching.
\*SMD is for the mean difference between groups divided by the pooled standard deviation.

**Figure 2.**
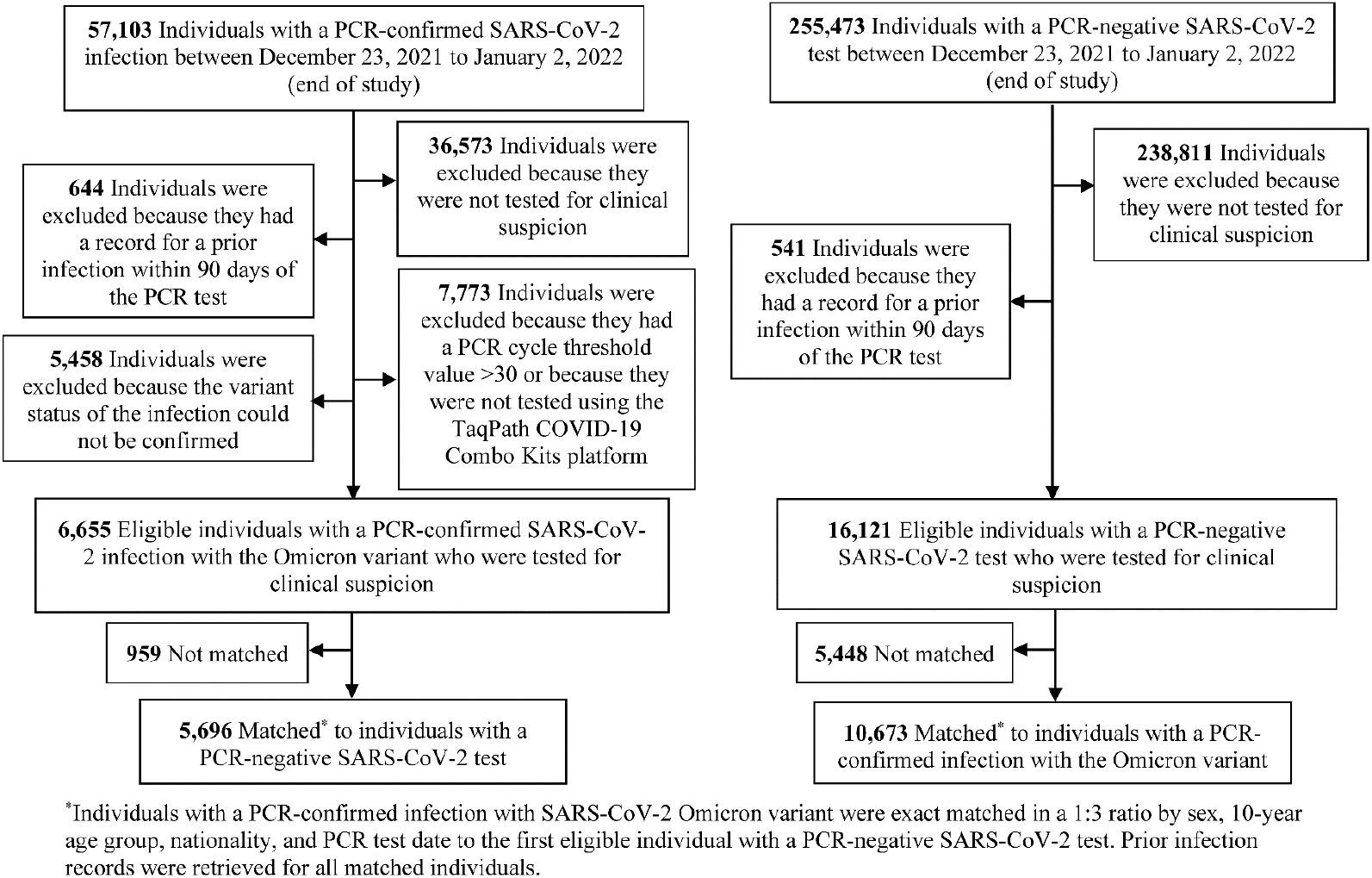
Flowchart describing the population selection process for investigating the effectiveness of prior infection in preventing reinfection with the Omicron variant.

Description of laboratory methods for the RT-qPCR testing and variant ascertainment are found in Section S1. All PCR testing was conducted at the Hamad Medical Corporation Central Laboratory or at Sidra Medicine Laboratory, following standardized protocols.

Matching of cases and controls was performed to control for known differences in the risk of exposure to SARS-CoV-2 infection in Qatar.^9,14–17^ Only cases with an RT-qPCR cycle threshold (Ct) value ≤30 and individuals tested because of clinical suspicion, that is presence of symptoms compatible with a respiratory tract infection, were included in analysis. These criteria were applied to ensure that *PES* is estimated against reinfections with at least some symptomatic disease and epidemiological relevance, as often reinfections occur with negligible symptoms and high Ct values, which are of less public health significance.^18^

Only the first PCR-positive test for a specific variant of interest was included for each case. A control was defined as the first PCR-negative test for any individual tested for clinical suspicion during the study period.^19–23^ Prior infection was defined as a PCR-confirmed infection ≥90 days before a new PCR-positive test.^7,24^ Individuals PCR-positive during the 90 days preceding the PCR test were therefore excluded from both cases and controls. These inclusion and exclusion criteria were implemented to minimize different types of potential bias, as informed by prior analyses.^20^

Each person who had a PCR-positive test result and hospital admission was subject to an infection severity assessment every three days until discharge or death, regardless of the length of the hospital stay or the time between the PCR-positive test and the final disease outcome.

Classification of COVID-19 case severity (acute-care hospitalization),^25^ criticality (intensive-care-unit (ICU) hospitalization),^25^ and fatality^26^ followed World Health Organization (WHO) guidelines, and assessments were made by trained medical personnel using individual chart reviews (Section S2).

The latter protocol for infection severity assessment was applied for Alpha, Beta, and Delta cases. However, with the recency of the Omicron epidemic wave, assessment of severity, criticality, and fatality of Omicron cases was completed for only a small number of cases. Therefore, only for Omicron cases, any acute-bed hospital admission associated with infection was used as a proxy for COVID-19 severity, and any ICU-bed hospital admission associated with infection was used as a proxy for COVID-19 criticality. Alpha, Beta, Delta, and Omicron cases that progressed to severe,^25^ critical,^25^ or fatal^26^ COVID-19 between the PCR-positive test result and the end of the study were classified based on their worst outcome, starting with death, followed by critical disease, and then severe disease.

Effectiveness of prior infection in preventing severe, critical, or fatal COVID-19 reinfection was also estimated, applying the same methodology. Here, cases (PCR-positive persons with a variant infection that progressed to a severe, critical, or fatal COVID-19) were exact matched to controls (PCR-negative persons) using the matching criteria specified above for each variant type.

The study was approved by the Hamad Medical Corporation and Weill Cornell Medicine-Qatar Institutional Review Boards with a waiver of informed consent. Reporting of the study followed STROBE guidelines (Table S1).

### Statistical analysis

All records of PCR testing in Qatar were examined for the selection of cases and controls and ascertainment of prior infection status. However, only matched samples of cases and controls were included in the analysis. Cases and controls were described using frequency distributions and measures of central tendency and compared using standardized mean differences (SMDs). SMD is defined as the difference in the mean of a covariate between groups divided by the pooled standard deviation, with SMD <0.1 indicating optimal balance across groups.

*PES* was derived as one minus the ratio of the odds of prior infection in cases (PCR-positive persons with variant infection), to the odds of prior infection in controls (PCR-negative persons):^6^

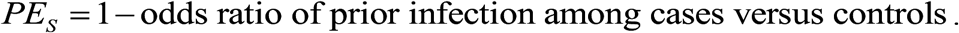

Odds ratios and associated 95% confidence intervals (CIs) were derived using conditional logistic regression, factoring the matching in the study design. This analytical approach minimizes potential bias that could arise due to variation in epidemic phase^4,27^ or other confounders.^9,14–17,28,29^ CIs were not adjusted for multiplicity. Interactions were not investigated.

Two sensitivity analyses were conducted to assess the robustness of estimates of *PES*. The first estimated *PES* by additionally adjusting for vaccination status in the conditional logistic regression. The second estimated *PES* after excluding all individuals with a record of vaccination prior to the PCR test used for defining cases and controls (Figures 3 and 4).

**Figure 3.**
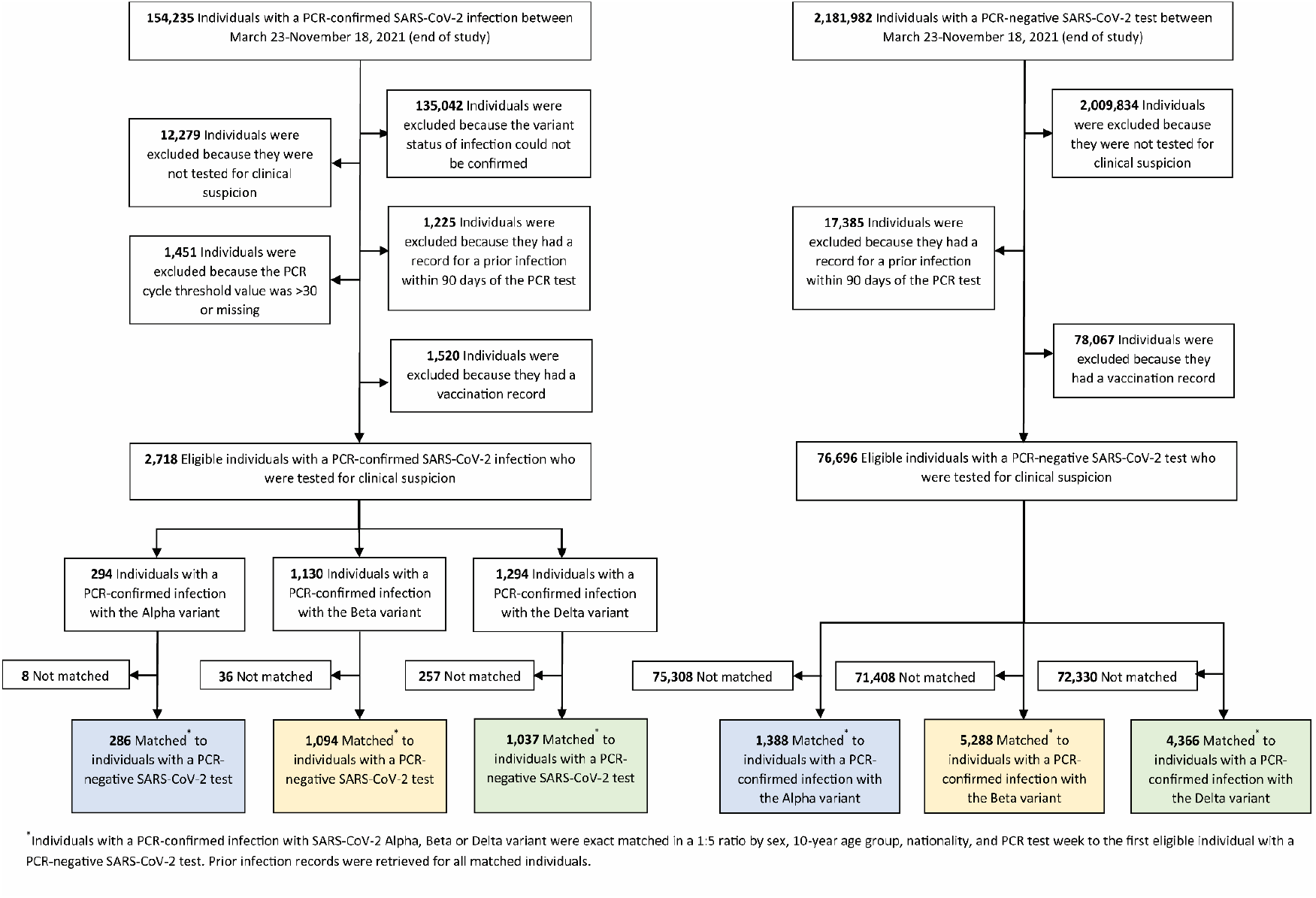
Flowchart describing the population selection process for the sensitivity analysis investigating the effectiveness of prior infection in preventing reinfection with the Alpha, Beta, and Delta variants after excluding those vaccinated.

**Figure 4.**
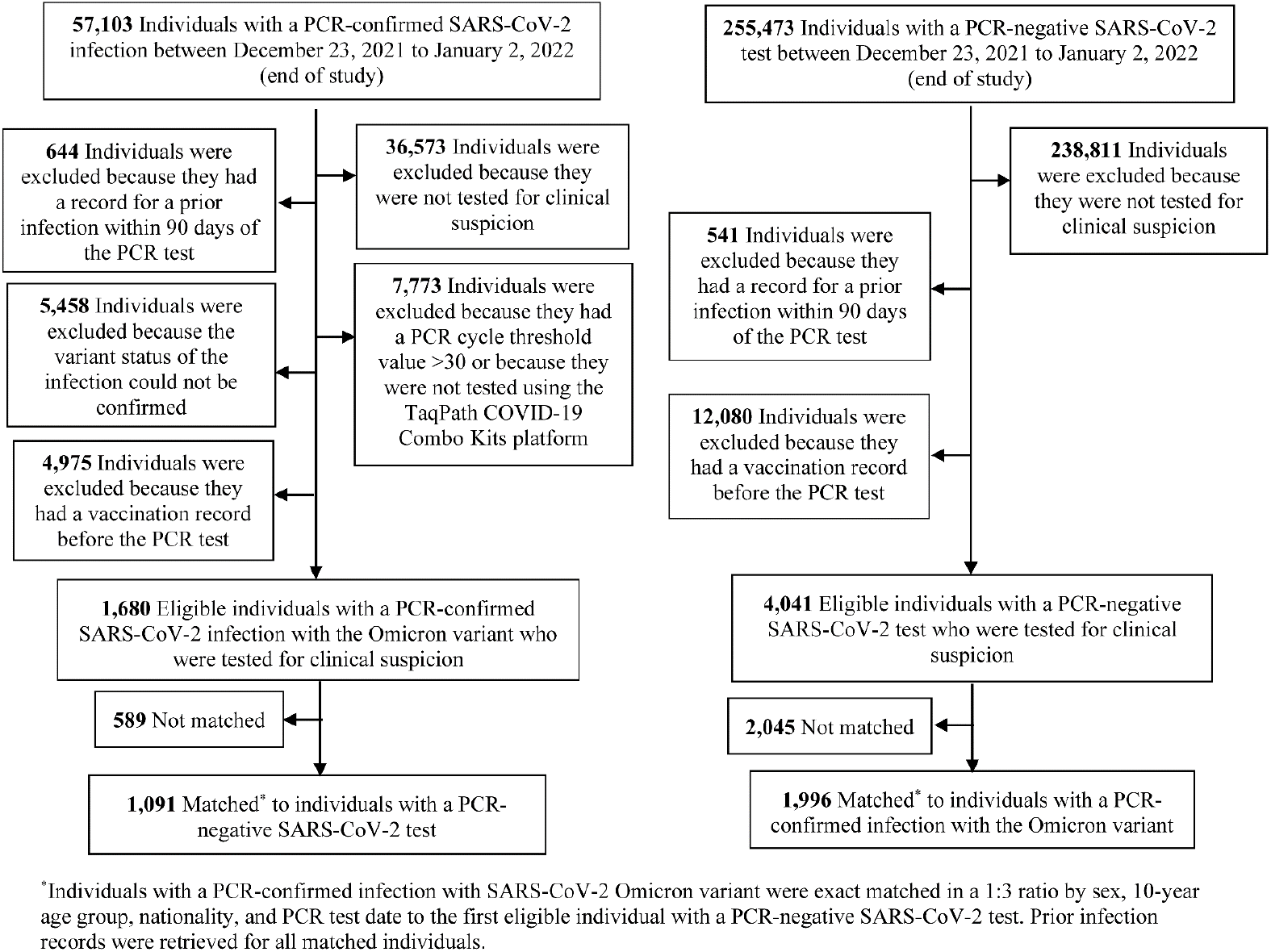
Flowchart describing the population selection process for the sensitivity analysis investigating the effectiveness of prior infection in preventing reinfection with the Omicron variant after excluding those vaccinated.

## Results

Figures 1–4 depict the selection of the study populations. Tables 1–2 show study population characteristics. The median time between prior infection and PCR test among cases and controls was 279 days (interquartile range (IQR), 194-313) for Alpha, 284 days (IQR, 210-313) for Beta, 253 days (IQR, 159-375) for Delta, and 314 days (IQR, 268-487) for Omicron.

Table 3 summarizes study results. *PES* against symptomatic reinfection was estimated at 90.2% (95% CI: 60.2-97.6) for Alpha, 84.8% (95% CI: 74.5-91.0) for Beta, 92.0% (95% CI: 87.9-94.7) for Delta, and 56.0% (95% CI: 50.6-60.9) for Omicron. Sensitivity analyses, adjusting for vaccination status or excluding vaccinated subjects from the analysis, confirmed the study results (Table 3), as expected for this study design, which is robust, irrespective of the approach employed to control for vaccine-induced immunity.^6^

**Table 3.** Effectiveness of SARS-CoV-2 prior infection against reinfection with Alpha, Beta, Delta, or Omicron variant.

|  | Cases (PCR-positive) |  | Controls (PCR-negative) |  | Effectiveness in %<br>(95% CI)<sup>‡</sup> |
| --- | --- | --- | --- | --- | --- |
|  | Prior infection | No prior infection | Prior infection | No prior infection |  |
| Effectiveness against symptomatic infection |  |  |  |  |  |
| A) Main analysis* |  |  |  |  |  |
| Alpha | 2 | 334 | 94 | 1,548 | 90.2 (60.2 to 97.6) |
| Beta | 15 | 1,321 | 450 | 6,084 | 84.8 (74.5 to 91.0) |
| Delta | 23 | 2,153 | 1,154 | 8,782 | 92.0 (87.9 to 94.7) |
| Omicron | 412 | 5,284 | 1,620 | 9,053 | 56.0 (50.6 to 60.9) |
| B) Adjusting for vaccination status in conditional logistic regression* |  |  |  |  |  |
| Alpha | 2 | 334 | 94 | 1,548 | 90.3 (60.4 to 97.6) |
| Beta | 15 | 1,321 | 450 | 6,084 | 84.0 (73.1 to 90.5) |
| Delta | 23 | 2,153 | 1,154 | 8,782 | 91.9 (87.8 to 94.7) |
| Omicron | 412 | 5,284 | 1,620 | 9,053 | 55.9 (50.5 to 60.8) |
| C) Excluding vaccinated individuals<sup>†</sup> |  |  |  |  |  |
| Alpha | 1 | 285 | 94 | 1,294 | 95.3 (66.0 to 99.3) |
| Beta | 11 | 1,084 | 312 | 4,976 | 83.9 (70.4 to 91.2) |
| Delta | 11 | 1,026 | 400 | 3,966 | 90.5 (81.9 to 94.6) |
| Omicron | 60 | 1,031 | 258 | 1,738 | 61.9 (48.2 to 72.0) |
| Effectiveness against severe, critical, or fatal COVID-19<sup>§</sup> |  |  |  |  |  |
| Alpha | 1 | 44 | 15 | 199 | 69.4 (-143.6 to 96.2) |
| Beta | 2 | 186 | 76 | 824 | 88.0 (50.7 to 97.1) |
| Delta | 0 | 135 | 56 | 528 | 100 (43.3 to 99.8)<sup>‡</sup> |
| Omicron | 2 | 70 | 39 | 167 | 87.8 (47.5 to 97.1) |

<sup>‡</sup>Cases and controls were exact matched one-to-five by sex, 10-year age group, nationality, and calendar week of PCR test in the Alpha, Beta, and Delta analyses (March 23-November 18, 2021; Figure 1), and one-to-three by sex, 10-year age group, nationality, and PCR test date in the Omicron analysis (December 23, 2021-Jan 2, 2022; Figure 2).
<sup>‡</sup>Cases and controls were exact matched one-to-five by sex, 10-year age group, nationality, and calendar week of PCR test in the Alpha, Beta, and Delta analyses (March 23-November 18, 2021; Figure 3), and one-to-three by sex, 10-year age group, nationality, and PCR test date in the Omicron analysis (December 23, 2021-Jan 2, 2022; Figure 4).
<sup>‡</sup>Effectiveness of prior infection in preventing reinfection was estimated using the test-negative, case-control study design.<sup>6</sup>
<sup>§</sup>Severity, criticality, and fatality of Alpha, Beta, and Delta cases were defined as per World Health Organization guidelines (Methods and Section S2). With the recency of the Omicron epidemic wave in Qatar, assessment of severity, criticality, and fatality of Omicron cases was completed for only a small number of cases. Therefore, any acute-bed hospital admission associated with Omicron infection was used as a proxy for COVID-19 severity. Any ICU-bed hospital admission associated with Omicron infection was used as a proxy for COVID-19 criticality.
<sup>¶</sup>The confidence interval could not be estimated using conditional logistic regression because of zero events among those with prior infection. Alternatively, the confidence interval was estimated using the standard error of the crude odds ratio after adding 0.5 to each number of cases with and without prior infection.

Only 1 Alpha, 2 Beta, 0 Delta, and 2 Omicron reinfections progressed to severe COVID-19 (Table 3). None progressed to critical or fatal COVID-19. *PES* against hospitalization or death due to reinfection was estimated at 69.4% (95% CI: −143.6-96.2) for Alpha, 88.0% (95% CI: 50.7-97.1) for Beta, 100% (95% CI: 43.3-99.8) for Delta, and 87.8% (95% CI: 47.5-97.1) for Omicron.

## Discussion

Protection afforded by prior infection in preventing symptomatic reinfection with Alpha, Beta, or Delta is robust, at about 90%, confirming earlier estimates.^1–3^ While such protection against reinfection with Omicron is lower, it is still considerable at nearly 60%. Prior-infection protection against hospitalization or death at reinfection appears robust, regardless of variant.

Individual-level data on co-morbid conditions were not available; therefore, they could not be explicitly factored into our analysis. However, only a small proportion of the study population may have had serious co-morbid conditions. Only 9% of the population of Qatar are ≥50 years of age,^8,9^ and 60% are young, expatriate craft and manual workers working in mega-development projects.^16,17,30^ The national list of persons prioritized to receive the vaccine during the first phase of vaccine roll-out included only 19,800 individuals of all age groups with serious co-morbid conditions. Matching of cases and controls on age may have indirectly and partially adjusted for presence of co-morbidities. With the young population of Qatar, our findings may not be generalizable to other countries where elderly citizens constitute a larger proportion of the total population.

*PES* was assessed using an observational, test-negative, case-control study design,^6^ rather than a cohort study design where individuals are followed up over time. However, the cohort study design applied in earlier analyses to estimate *PES* in the same population of Qatar yielded findings similar to those of the test-negative case-control design,^1,2,6,7,31^ supporting the validity of this design in estimating *PES*. It even appears that the test-negative study design may be less susceptible to some forms of bias than the cohort study design.^6^

Nonetheless, one cannot exclude the possibility that in real-world data, bias could arise in unexpected ways, or from unknown sources, such as subtle differences in test-seeking behavior or changes in the pattern of testing with introduction of other testing modalities, such as rapid antigen testing.

Notwithstanding these limitations, consistent findings were reached in both the main and sensitivity analyses. Estimates for the effectiveness of prior infection against reinfection with the Alpha and Beta variants were also consistent and similar to those generated earlier in the same population of Qatar using cohort study designs.^1,2^

## Data Availability

The dataset of this study is a property of the Qatar Ministry of Public Health that was provided to the researchers through a restricted-access agreement that prevents sharing the dataset with a third party or publicly. Future access to this dataset can be considered through a direct application for data access to Her Excellency the Minister of Public Health (https://www.moph.gov.qa/english/Pages/default.aspx). Aggregate data are available within the manuscript and its Supplementary information.

## Acknowledgements

We acknowledge the many dedicated individuals at Hamad Medical Corporation, the Ministry of Public Health, the Primary Health Care Corporation, the Qatar Biobank, Sidra Medicine, and Weill Cornell Medicine – Qatar for their diligent efforts and contributions to make this study possible.

The authors are grateful for support from the Biomedical Research Program and the Biostatistics, Epidemiology, and Biomathematics Research Core, both at Weill Cornell Medicine-Qatar, as well as for support provided by the Ministry of Public Health, Hamad Medical Corporation, and Sidra Medicine. The authors are also grateful for the Qatar Genome Programme and Qatar University Biomedical Research Center for institutional support for the reagents needed for the viral genome sequencing. Statements made herein are solely the responsibility of the authors.

The funders of the study had no role in study design, data collection, data analysis, data interpretation, or writing of the article.

## Author contributions

HNA and HC co-designed the study, performed the statistical analyses, and co-wrote the first draft of the article. LJA conceived and co-designed the study, led the statistical analyses, and co-wrote the first draft of the article. PT and MRH conducted the multiplex, RT-qPCR variant screening and viral genome sequencing. HY, FMB, and HAK conducted viral genome sequencing. All authors contributed to data collection and acquisition, database development, discussion and interpretation of the results, and to the writing of the manuscript. All authors have read and approved the final manuscript.

## Supplementary Appendix

### Section S1. Laboratory methods

#### Real-time reverse-transcription polymerase chain reaction testing

Nasopharyngeal and/or oropharyngeal swabs were collected for polymerase chain reaction (PCR) testing and placed in Universal Transport Medium (UTM). Aliquots of UTM were: extracted on a QIAsymphony platform (QIAGEN, USA) and tested with real-time reverse-transcription PCR (RT-qPCR) using TaqPath COVID-19 Combo Kits (Thermo Fisher Scientific, USA) on an ABI 7500 FAST (Thermo Fisher, USA); tested directly on the Cepheid GeneXpert system using the Xpert Xpress SARS-CoV-2 (Cepheid, USA); or loaded directly into a Roche cobas 6800 system and assayed with a cobas SARS-CoV-2 Test (Roche, Switzerland). The first assay targets the viral S, N, and ORF1ab gene regions. The second targets the viral N and E-gene regions, and the third targets the ORF1ab and E-gene regions.

All PCR testing was conducted at the Hamad Medical Corporation Central Laboratory or Sidra Medicine Laboratory, following standardized protocols.

#### Classification of infections by variant type

Surveillance for SARS-CoV-2 variants in Qatar is mainly based on viral genome sequencing and multiplex RT-qPCR variant screening^1^ of random positive clinical samples,^2–7^ complemented by deep sequencing of wastewater samples.^4,8^

Between March 23, 2021 and November 18, 2021 (prior to suspected introduction of the Omicron variant), RT-qPCR genotyping of 19,234 randomly collected SARS-CoV-2-positive specimens on a weekly basis identified 3,494 (18.2%) Alpha (B.1.1.7)-like cases, 5,768 (30.0%)

Beta (B.1.351)-like cases, 9,914 (51.5%) “other” variant cases, and 58 (0.3%) B.1.375-like or B.1.258-like cases.^4,6^

The accuracy of the RT-qPCR genotyping was verified against either Sanger sequencing of the receptor-binding domain (RBD) of SARS-CoV-2 surface glycoprotein (S) gene, or by viral whole-genome sequencing on a Nanopore GridION sequencing device. From 236 random samples (27 Alpha-like, 186 Beta-like, and 23 “other” variants), PCR genotyping results for Alpha-like, Beta-like, and ‘other’ variants were in 88.8% (23 out of 27), 99.5% (185 out of 186), and 100% (23 out of 23) agreement with the SARS-CoV-2 lineages assigned by sequencing.

Within the “other” variant category, Sanger sequencing and/or Illumina sequencing of the RBD of SARS-CoV-2 spike gene on 728 random samples confirmed that 701 (96.3%) were Delta cases and 17 (2.3%) were other variant cases, with 10 (1.4%) samples failing lineage assignment.^6,8^ Accordingly, a Delta case was proxied as any “other” case identified through the RT-qPCR based variant screening.

All the variant RT-qPCR screening was conducted at the Sidra Medicine Laboratory following standardized protocols.

Surveillance for Omicron infection was performed using the TaqPath COVID-19 Combo Kit platform (Thermo Fisher Scientific, USA^9^) applying the criterion of an RT-qPCR Ct value ≤30 for both the N and ORF1ab genes, but a negative outcome for the S gene (S-gene “target failure”).

### Section S2. COVID-19 severity, criticality, and fatality classification

Severe Coronavirus Disease 2019 (COVID-19) disease was defined per the World health Organization (WHO) classification as a severe acute respiratory syndrome coronavirus 2 (SARS-CoV-2) infected person with “oxygen saturation of <90% on room air, and/or respiratory rate of >30 breaths/minute in adults and children >5 years old (or ≥60 breaths/minute in children <2 months old or ≥50 breaths/minute in children 2-11 months old or ≥40 breaths/minute in children 1–5 years old), and/or signs of severe respiratory distress (accessory muscle use and inability to complete full sentences, and, in children, very severe chest wall indrawing, grunting, central cyanosis, or presence of any other general danger signs)”.^10^ Detailed WHO criteria for classifying SARS-CoV-2 infection severity can be found in the WHO technical report.^10^

Critical COVID-19 disease was defined per WHO classification as a SARS-CoV-2 infected person with “acute respiratory distress syndrome, sepsis, septic shock, or other conditions that would normally require the provision of life sustaining therapies such as mechanical ventilation (invasive or non-invasive) or vasopressor therapy”.^10^ Detailed WHO criteria for classifying SARS-CoV-2 infection criticality can be found in the WHO technical report.^10^

COVID-19 death was defined per WHO classification as “a death resulting from a clinically compatible illness, in a probable or confirmed COVID-19 case, unless there is a clear alternative cause of death that cannot be related to COVID-19 disease (e.g. trauma). There should be no period of complete recovery from COVID-19 between illness and death. A death due to COVID-19 may not be attributed to another disease (e.g. cancer) and should be counted independently of preexisting conditions that are suspected of triggering a severe course of COVID-19”. Detailed WHO criteria for classifying COVID-19 death can be found in the WHO technical report.^11^

**Table S1.** STROBE checklist for case-control studies.

|  | Item No | Recommendation | Main text |
| --- | --- | --- | --- |
| Title and abstract | 1 | (a) Indicate the study’s design with a commonly used term in the title or the abstract | Abstract |
|  |  | (b) Provide in the abstract an informative and balanced summary of what was done and what was found | Abstract |
| Introduction |  |  |  |
| Background/rationale | 2 | Explain the scientific background and rationale for the investigation being reported | Introduction |
| Objectives | 3 | State specific objectives, including any prespecified hypotheses | Introduction |
| Methods |  |  |  |
| Study design | 4 | Present key elements of study design | Methods (‘Study population, data sources, and study design’) |
| Setting | 5 | Describe the setting, locations, and relevant dates, including periods of recruitment, exposure, follow-up, and data collection | Methods (‘Study population, data sources, and study design’) & Figures 1-4 |
| Participants | 6 | (a) Give the eligibility criteria, and the sources and methods of case ascertainment and control selection. Give the rationale for the choice of cases and controls | Methods (‘Study population, data sources, and study design’) & Figures 1-4 |
|  |  | (b) For matched studies, give matching criteria and the number of controls per case |  |
| Variables | 7 | Clearly define all outcomes, exposures, predictors, potential confounders, and effect modifiers. Give diagnostic criteria, if applicable | Methods (‘Study population, data sources, and study design’ & ‘Statistical analysis’), Supp. Sections S1 & S2 |
| Data sources/measurement | 8 | For each variable of interest, give sources of data and details of methods of assessment (measurement). Describe comparability of assessment methods if there is more than one group | Methods (‘Study population, data sources, and study design’ & ‘Statistical analysis’) & Tables 1-2 |
| Bias | 9 | Describe any efforts to address potential sources of bias | Methods (‘Study population, data sources, and study design’ & ‘Statistical analysis’), & Figures 3-4 |
| Study size | 10 | Explain how the study size was arrived at | Methods (‘Study population, data sources, and study design’) & Figures 1-2 |
| Quantitative variables | 11 | Explain how quantitative variables were handled in the analyses. If applicable, describe which groupings were chosen and why | Methods (‘Study population, data sources, and study design’ & ‘Statistical analysis’) & Tables 1-2 |
| Statistical methods | 12 | (a) Describe all statistical methods, including those used to control for confounding | Methods (‘Statistical analysis’) & Table 3 |
|  |  | (b) Describe any methods used to examine subgroups and interactions | Methods (‘Statistical analysis’), Figures 3-4, & Table 1 |
|  |  | (c) Explain how missing data were addressed | NA, see Methods (‘Study population, data sources, and study design’) |
|  |  | (d) If applicable, explain how matching of cases and controls was addressed | Methods (‘Study population, data sources, and study design’) & Tables 1-2 |
|  |  | (e) Describe any sensitivity analyses | Methods (‘Statistical analysis’), Figures 3-4, & Table 3 |
| Results |  |  |  |
| Participants | 13 | (a) Report numbers of individuals at each stage of study—eg numbers potentially eligible, examined for eligibility, confirmed eligible, included in the study, completing follow-up, and analysed | Figures 1-2 |
|  |  | (b) Give reasons for non-participation at each stage |  |
|  |  | (c) Consider use of a flow diagram |  |
| Descriptive data | 14 | (a) Give characteristics of study participants (eg demographic, clinical, social) and information on exposures and potential confounders | Tables 1-2 |
|  |  | (b) Indicate number of participants with missing data for each variable of interest | NA, see Methods (‘Study population, data sources, and study design’) |
| Outcome data | 15 | Report numbers in each exposure category, or summary measures of exposure | Results & Table 3 |
| Main results | 16 | (a) Give unadjusted estimates and, if applicable, confounder-adjusted estimates and their precision (eg, 95% confidence interval). Make clear which confounders were adjusted for and why they were included | Results & Table 3 |
|  |  | (b) Report category boundaries when continuous variables were categorized | Tables 1-2 |
|  |  | (c) If relevant, consider translating estimates of relative risk into absolute risk for a meaningful time period | NA |
| Other analyses | 17 | Report other analyses done—eg analyses of subgroups and interactions, and sensitivity analyses | Results & Table 3 |
| **Discussion** |  |  |  |
| Key results | 18 | Summarise key results with reference to study objectives | Discussion, paragraph 1 |
| Limitations | 19 | Discuss limitations of the study, taking into account sources of potential bias or imprecision. Discuss both direction and magnitude of any potential bias | Discussion paragraphs 2-4 |
| Interpretation | 20 | Give a cautious overall interpretation of results considering objectives, limitations, multiplicity of analyses, results from similar studies, and other relevant evidence | Discussion paragraph 5 |
| Generalisability | 21 | Discuss the generalisability (external validity) of the study results | Discussion paragraphs 2-4 |
| **Other information** |  |  |  |
| Funding | 22 | Give the source of funding and the role of the funders for the present study and, if applicable, for the original study on which the present article is based | Acknowledgements |

Abbreviations: NA, not applicable; p. page; Supp. Supplementary Appendix.

